# Comparative Evaluation of Advanced AI Reasoning Models in Pediatric Clinical Decision Support: ChatGPT O1 vs. DeepSeek-R1

**DOI:** 10.1101/2025.01.27.25321169

**Authors:** Gianluca Mondillo, Simone Colosimo, Alessandra Perrotta, Vittoria Frattolillo, Mariapia Masino

**Author notes:** Address correspondence to: Gianluca Mondillo, Department of Woman, Child and of General and Specialized Surgery, Università degli Studi della Campania “Luigi Vanvitelli”, Via Luigi De Crecchio 4, 80138, Napoli, Italy, 0039-3391542528.

## Abstract

**Introduction:** The adoption of advanced reasoning models, such as ChatGPT O1 and DeepSeek-R1, represents a pivotal step forward in clinical decision support, particularly in pediatrics. ChatGPT O1 employs “chain-of-thought reasoning” (CoT) to enhance structured problem-solving, while DeepSeek-R1 introduces self-reflection capabilities through reinforcement learning. This study aimed to evaluate the diagnostic accuracy and clinical utility of these models in pediatric scenarios using the MedQA dataset.

**Materials and Methods:** A total of 500 multiple-choice pediatric questions from the MedQA dataset were presented to ChatGPT O1 and DeepSeek-R1. Each question included four or more options, with one correct answer. The models were evaluated under uniform conditions, with performance metrics including accuracy, Cohen’s Kappa, and chi-square tests applied to assess agreement and statistical significance. Responses were analyzed to determine the models effectiveness in addressing clinical questions.

**Results:** ChatGPT O1 achieved a diagnostic accuracy of 92.8%, significantly outperforming DeepSeek-R1, which scored 87.0% (*p <* 0.00001). The CoT reasoning technique used by ChatGPT O1 allowed for more structured and reliable responses, reducing the risk of errors. Conversely, DeepSeek-R1, while slightly less accurate, demonstrated superior accessibility and adaptability due to its open-source nature and emerging self-reflection capabilities. Cohen’s Kappa (K=0.20) indicated low agreement between the models, reflecting their distinct reasoning strategies.

**Conclusions:** This study highlights the strengths of ChatGPT O1 in providing accurate and coherent clinical reasoning, making it highly suitable for critical pediatric scenarios.

DeepSeek-R1, with its flexibility and accessibility, remains a valuable tool in resource-limited settings. Combining these models in an ensemble system could leverage their complementary strengths, optimizing decision support in diverse clinical contexts. Further research is warranted to explore their integration into multidisciplinary care teams and their application in real-world clinical settings.

## Introduction

Advanced artificial intelligence (AI) models have rapidly transformed various sectors, including medicine, by enabling the management and analysis of vast amounts of data. Among these, reasoning models stand out for their ability to address complex problems through structured and sequential thinking processes, marking a significant evolution from traditional Large Language Models (LLMs) [1]. A significant example of this evolution is represented by the ChatGPT O1 (OpenAI, September 2024) [2] and DeepSeek-R1 (DeepSeek, January 2025) [3] models. ChatGPT O1 stands out for implementing the chain-of-thought reasoning (CoT) methodology [4], which enables the model to break down complex problems into a series of sequential logical steps, improving consistency and reducing the risk of incorrect or incomplete answers. On the other hand, DeepSeek-R1, an open-source model based on reinforcement learning techniques, offers emerging self-reflection capabilities, making it a valuable resource for medical applications in low-resource settings, thanks to its accessibility and flexibility. These capabilities are particularly promising in pediatrics, a field where the accuracy and timeliness of clinical decisions are critical. However, the use of reasoning models in medicine requires careful evaluation of their performance, especially regarding their ability to balance sensitivity and specificity in critical clinical scenarios. In this study, we compared the performance of the ChatGPT O1 and DeepSeek-R1 models on a sample of pediatric questions drawn from the MedQA dataset [5,6], aiming to identify their potential and limitations in clinical application.

## Materials and Methods

The study utilized the MedQA dataset, a standard resource for evaluating LLMs in medicine. The dataset includes multiple-choice questions on various medical topics, validated by experts, including pediatrics. To ensure adequate representation, 500 questions concerning pediatric patients (ages 0–16 years, median 7 years) were randomly selected. Each question presented four or more answer options, with only one correct answer. The models analyzed in the study were:

1. ChatGPT O1: An advanced reasoning model implementing chain-of-thought reasoning to enhance structured reasoning. Access to the model was obtained through a monthly subscription, with a limit of 50 weekly messages [7].
2. DeepSeek-R1: An open-source reasoning model based on reinforcement learning, designed to generate reasoning capabilities without supervision. The free version used supports up to 50 daily messages [8].

Both models were presented with the same questions as a single prompt, without additional instructions. To comply with the weekly limit of 50 messages imposed by the ChatGPT O1 model, the questions were delivered in blocks of 25 questions at a time, totaling 20 test sessions per model. This division allowed for the completion of the entire question set without exceeding the imposed weekly limits, maintaining uniform test conditions between the models. The answers provided by the models were compared to the correct ones in the MedQA dataset.

Model performance was evaluated using the following metrics:

- Accuracy: Percentage of correct answers out of the total.
- Cohen’s Kappa: Agreement between the models, accounting for chance agreement.
- Chi-square: Statistical test to assess the significance of differences in the classification patterns of the models.

### Statistical Analysis

Data analysis was performed using Python (SciPy 1.14.1) to calculate the metrics and statistical tests. A p-value of *<* 0.05 was considered significant.

## Results

The analysis of the responses from the ChatGPT O1 and DeepSeek-R1 models revealed significant differences in their performance. The ChatGPT O1 model achieved an accuracy of 92.8% (464 correct answers out of 500), while DeepSeek-R1 achieved an accuracy of 87.0% (435 correct answers out of 500) (Figure 1). These results indicate a superior ability of ChatGPT O1 to provide correct answers overall.

**Figure 1.**
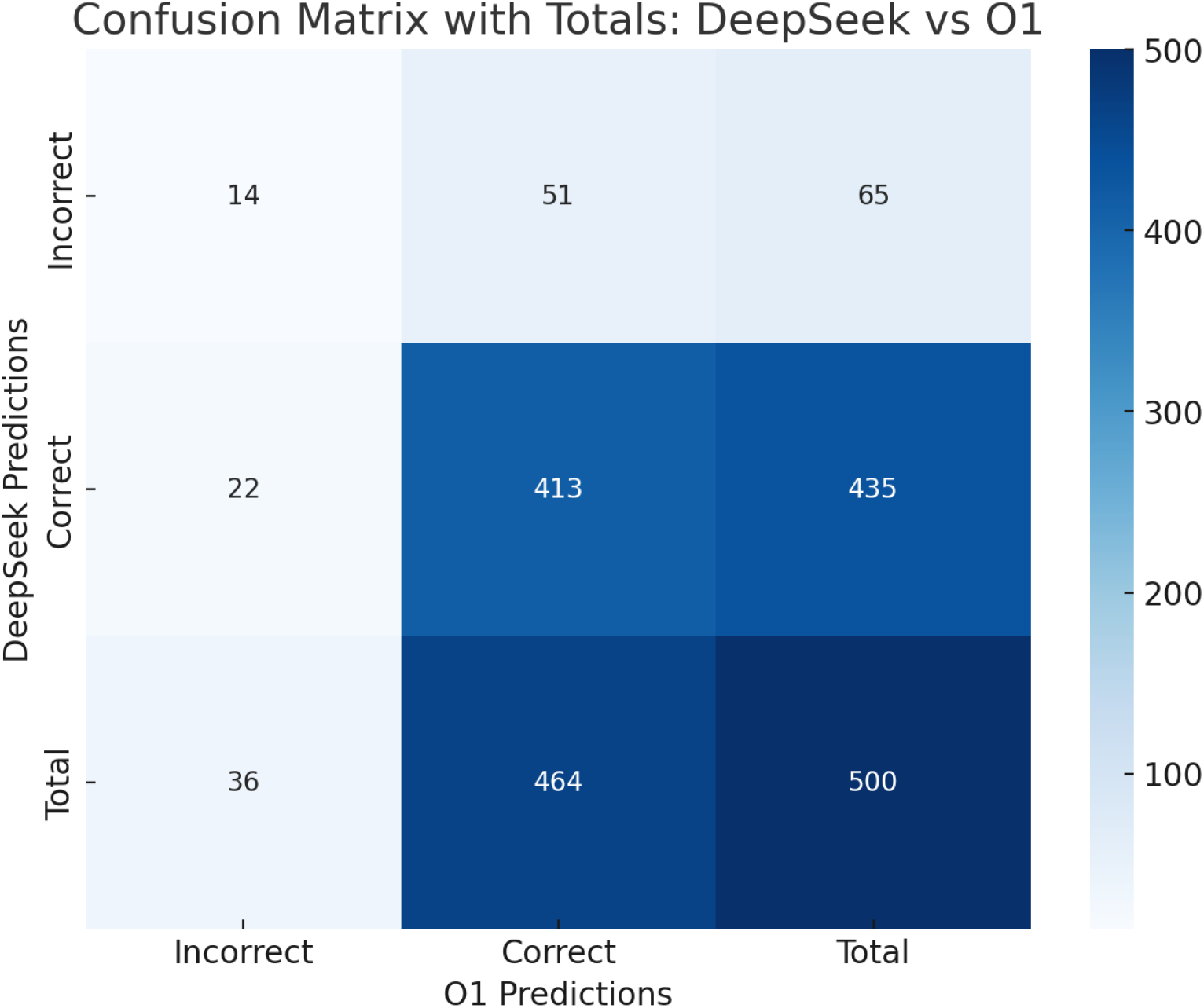
Confusion Matrices of the analyzed models.

Cohen’s Kappa (K=0.20) revealed a low agreement between the models, indicating that their predictions are not overlapping and may reflect different reasoning strategies. Additionally, the chi-square test with Yates correction (*p <* 0.00001) confirmed that the differences in the classification patterns of the two models are statistically significant (Table 1).

**Table 1.** Comparisons of correct response rates among models.

| Models Comparisons | Correct Answers Model 1 | Correct Answers Model 2 | p-value |
| --- | --- | --- | --- |
| ChatGPT O1 (Model 1) vs DeepSeek R1 (Model 2) | 92.8% (464/500) | 87.0% (435/500) | p<0.00001 |

## Discussion

The comparative analysis between the ChatGPT O1 and DeepSeek-R1 models highlighted differences in performance and design principles. The two models demonstrate distinctive approaches reflected in accuracy metrics and clinical application potential. The ChatGPT O1 model stands out for its overall accuracy of 92.8%, higher than the 87.0% of DeepSeek-R1, demonstrating greater reliability in providing correct answers. This characteristic makes ChatGPT O1 particularly suitable in clinical contexts where minimizing diagnostic errors is paramount. For example, in critical scenarios such as managing a newborn with signs of sepsis, a model like ChatGPT O1 could provide more reliable answers, reducing the risk of severe clinical consequences. This result may be attributed to the adoption of chain-of-thought reasoning, a technique that allows the model to address complex problems by breaking them into sequential steps, thereby enhancing structured reasoning. However, the model’s accessibility is limited by significant practical restrictions, such as the need for a paid subscription allowing only 50 weekly messages. These barriers could be a hindrance in resource-limited contexts, especially during intensive educational activities or extensive clinical simulations. On the other hand, DeepSeek-R1, while achieving slightly lower accuracy, emerges as an accessible and innovative solution due to its entirely open-source nature. This feature makes it particularly useful in healthcare settings with limited resources or for academic projects requiring free and flexible tools. DeepSeek-R1 employs a reinforcement learning-based (RL) approach during pretraining, enabling the model to develop advanced reasoning capabilities without relying on traditional supervised pretraining. A distinctive feature of DeepSeek-R1 is its emerging self-reflection capability, defined as self-evolution, through which the model autonomously verifies and optimizes its logical steps, improving its performance on complex tasks. This functionality could be particularly useful in complex queries, such as “What are the next steps in managing a child with suspected viral encephalitis?” where multi-level analysis is necessary. These different approaches may explain the low level of agreement calculated. However, the model has documented limitations, such as sensitivity to few-shot prompts, which could compromise performance in less structured contexts, and language mixing, with parts of the responses generated in English and others in Chinese, potentially posing a significant barrier in exclusively English-speaking contexts or scenarios requiring strictly monolingual communication [9]. In our study, the model always provided responses in English. From a technical perspective, the two models offer distinctive contributions: ChatGPT O1 is designed to maximize structured reasoning through the implementation of advanced techniques like chain-of-thought reasoning, making it particularly suitable for complex clinical contexts. Conversely, DeepSeek-R1 stands out for its flexibility and free availability, features that make it more accessible in resource-limited scenarios. The divergence in results, confirmed by the chi-square test (*p <* 0.00001), suggests that the differences between the two models are not attributable to chance. This finding reflects significantly different design choices that could be leveraged to develop an integrated approach, such as an ensemble system. An ensemble system, such as a Multi-Agent AI, could combine the strengths of the two models: the sensitivity and accuracy of ChatGPT O1, essential for minimizing the risk of diagnostic errors in critical cases, with the flexibility and accessibility of DeepSeek, ideal for ensuring operational continuity and solid performance even in less structured contexts. For example, in a pediatric decision-support scenario, the system could delegate complex and high-risk case analyses to ChatGPT O1, while DeepSeek could be utilized for direct questions and repetitive processes, ensuring greater overall efficiency. This integrated approach would allow balancing sensitivity, precision, and accessibility, offering more reliable and adaptable decision support to different clinical needs. Future studies could focus on the development of such a system to optimize overall performance through a synergistic integration of the two architectures.

## Conclusion

This study highlighted the fundamental differences between the ChatGPT O1 and DeepSeek-R1 models, each characterized by specific strengths. ChatGPT O1 proved particularly effective in complex reasoning, making it ideal for critical clinical situations. DeepSeek-R1, on the other hand, stands out for its accessibility and flexibility, representing a valuable solution in resource-limited settings. These findings underscore how the models, despite adopting different approaches, can both contribute to improving decision support in pediatrics, adapting to different needs and clinical scenarios.

## Data Availability

All data produced in the present study are available upon reasonable request to the authors

## Acknowledgments

We would like to thank the developers of the MedQA dataset for making their data publicly available. We also appreciate the support provided by OpenAI and DeepSeek in making their respective models accessible for research purposes.

## Declarations

### Ethical approval

Not applicable.

### Informed Consent

Not applicable.

### Funding source

No funding was secured for this study.

### Competing interests

The authors have no conflicts of interest to disclose.

### Clinical trial number

Not applicable.

## Notes

### Competing Interest Statement

The authors have declared no competing interest.

### Funding Statement

This study did not receive any funding

